# Clinical evaluation of a fully automated, lab developed multiplex RT-PCR assay integrating dual-target SARS-CoV-2 and Influenza–A/B detection on a high-throughput platform

**DOI:** 10.1101/2020.10.25.20215285

**Authors:** Dominik Nörz, Armin Hoffmann, Martin Aepfelbacher, Susanne Pfefferle, Marc Lütgehetmann

## Abstract

1

**Background:** Laboratories worldwide face high demands for molecular testing during the SARS-CoV-2 pandemic that might be further aggravated with the upcoming influenza season in the northern hemisphere. Considering that symptoms of influenza are largely undistinguishable from COVID-19, both SARS-CoV-2 and the Influenza viruses require concurrent testing by RT-PCR in patients presenting with symptoms of respiratory tract infection. In this study, we adapted and evaluated a laboratory developed multiplex RT-PCR assay for simultaneous detection of SARS-CoV-2 (dual-target), Influenza-A and Influenza-B (SC2/InflA/InflB-UCT) on a fully automated high-throughput system (cobas6800).

**Methods:** Analytical performance was assessed by serial dilution of quantified reference material and cell culture stocks in transport medium, including pre-treatment for chemical inactivation. For clinical evaluation, residual portions of 164 predetermined patient samples containing SARS-CoV-2 (n=52), Influenza-A (n=43) or Influenza-B (n=19), as well as a set of negative samples was subjected to the novel multiplex assay.

**Results:** The assay demonstrated analytical performance comparable to currently available commercial tests, with limits of detection of 94.9 cp/ml for SARS-CoV-2, 14.6 cp/ml for Influenza-A and 422.3 cp/ml for Influenza-B. Clinical evaluation showed excellent agreement with the comparator assays (sensitivity 98.1%, 97.7% and 100% for Sars-CoV-2, Influenza-A and -B respectively).

**Conclusion:** The SC2/InflA/InflB-UCT allows for efficient high-throughput testing for all three pathogens and thus provides streamlined diagnostics while conserving resources during the Influenza-season.

**Highlights:**

- Simultaneous detection of highly pathogenic respiratory viruses Influenza-A/B and SARS-CoV-2
- Including a dual-target assay for SARS-CoV-2 detection
- Full automation on the cobas6800 high-throughput platform

## 2 Introduction

The upcoming Influenza-season of 2020/21 will further aggravate the strain on diagnostic laboratories, already facing unprecedented demand for molecular diagnostics due to the ongoing SARS-CoV-2 pandemic. Just like COVID-19, Influenza is a major concern for infection control within healthcare facilities and symptoms are largely indistinguishable, particularly in the early phase of disease (1, 2). Consequently, SARS-CoV-2 and the Influenza viruses need to be concurrently tested for by RT-PCR before contact precaution measures can be lifted for symptomatic patients. In light of the continuing worldwide shortage of supplies for nucleic acid extraction and PCR diagnostics, it appears desirable to be able to screen for all three viruses (SARS-CoV-2, Influenza-A and Influenza-B) within the same reaction.

The cobas6800 system is a fully automated sample-to-result high-throughput platform, requiring minimal hands-on-time and able to perform up to 384-tests in an 8-hour shift. The instrument was previously evaluated for the detection of Influenza viruses in respiratory swabs (3) and is currently seeing increasing use for automated SARS-CoV-2 diagnostics (4, 5). The aim of this study was to establish and evaluate a multiplex assay for the detection of SARS-CoV-2, Influenza-A and Influenza-B on the open mode of the cobas6800 system (cobas omni utility channel).

## 3 Materials and Methods

### 3.1 SC2/InflA/InflB-UCT setup and preparation

A set of published RT-PCR assays for SARS-CoV-2, Influenza-A- and Influenza-B virus was selected and adapted for use on the cobas6800 system, see *table 1* (6-10). Primers were modified with 2’O-methylated RNA-bases at their penultimate positions to reduce formation of primer dimers. Double-quenched probes were used to lower background-fluorescence. All primers and probes were tested for contaminations prior to use, in particular concerning material reactive for SARS-CoV-2 in custom made commercial primers.

**Table 1:**
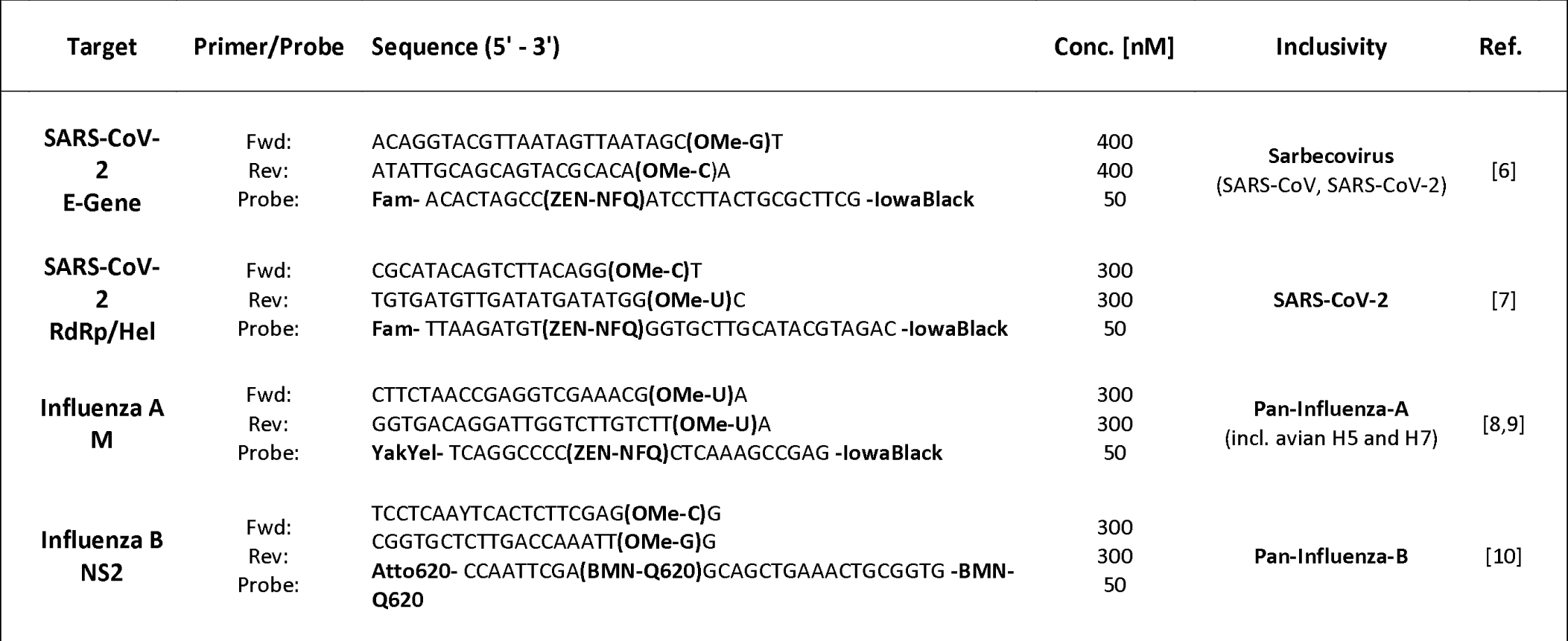
Assays used for the SC2/InflA/InflB-UCT. Primers and probes were custom made and procured from Integrated DNA Technologies (Illinois, USA), Biomers.net GmbH (Ulm, Germany) and Ella Biotech GmbH (Martinsried, Germany). OMe, 2’O-methyl RNA; NFQ, Non-fluorescent quencher; YakYel, Yakima Yellow

Primers and probes were diluted in MMX-R2-reagent to final concentrations as indicated above (*table 1*) to form the MMR2-Mastermix (MMRX-R2-reagent already contains the internal control assay by default). 6 ml of MMR2-Mastermix were loaded into cobas omni utility channel cassettes according to instructions by the manufacturer. The run profile for the SC2/InflA/InflB-UCT multiplex assay was configured using the cobas omni Utility Channel Software as indicated in *table 2*.

**Table 2:** Run-profile for the SC2/InflA/InflB, set up using the cobas omni utility channel tool.

| Software settings |  |  |  |  |  |
| --- | --- | --- | --- | --- | --- |
| Sample type | Alcohol Based Sample (400 µL) |  |  |  |  |
| Channels | 1: (Not assigned) | 2: SC2 | 3: InflA | 4: InflB | 5: IC |
| RFI |  | 1.25 | 1.25 | 1.25 | 1.15 |
| PCR cycling conditions |  |  |  |  |  |
|  | UNG incubation | Pre-PCR step | 1<sup>st</sup> measurement | 2<sup>nd</sup> measurement | Cooling |
| No. of cycles | Predefined | 1 | 5 | 45 | Predefined |
| No. of steps |  | 3 | 2 | 2 |  |
| Temperature |  | 55°C; 60°C; 65°C | 95°C; 55°C | 91°C; 58°C |  |
| Hold time |  | 120 s; 360 s; 240 s | 5 s; 30 s | 5 s; 25 s |  |
| Data acquisition |  | None | End of each cycle | End of each cycle |  |

### 3.2 Limit of detection (LoD), linearity and cross-reactivity

Analytical limit of detection (LoD) was determined for all three targets simultaneously by serial dilution of reference material in Amies medium including cobas PCR Media (1:1) as matrix. For SARS-CoV-2, a stock of cell culture supernatant containing SARS-CoV-2 (11) was quantified using the SARS-CoV-2 IVD Test for the cobas6800 (12) with the Qnostics “SARS-CoV-2 Q Control 01” as reference for quantification. For Influenza-A and Influenza-B, reference material was acquired from Qnostics (“INFA Medium Q Control 01” and “INFB Medium Q Control 01”) and used directly for LoD experiments. A total of 8 different concentrations was tested with 8 repeats each. (SC2 [cp/ml]-InflA [cp/ml]-InflB [cp/ml], 1000-1000-2000, 333-333-666, 100-100-200, 50-50-100, 25-25-50, 10-10-20, 3-3-6, 1-1-2) Linearity was determined for all targets simultaneously by 10-fold serial dilution, 5 repeats each step, using cell culture supernatant and Vaxigrip Tetravalent Influenza vaccine (Sanofi Pasteur, France) to spike Amies medium including cobas PCR Media (1:1) (*Figure 1*). Inclusivity and cross-reactivity were verified using external quality control panels by INSTAND e.V. (Düsseldorf, Germany) and clinical samples containing a variety of respiratory pathogens, including endemic human coronaviruses (see *table 4*).

**Table 3:**
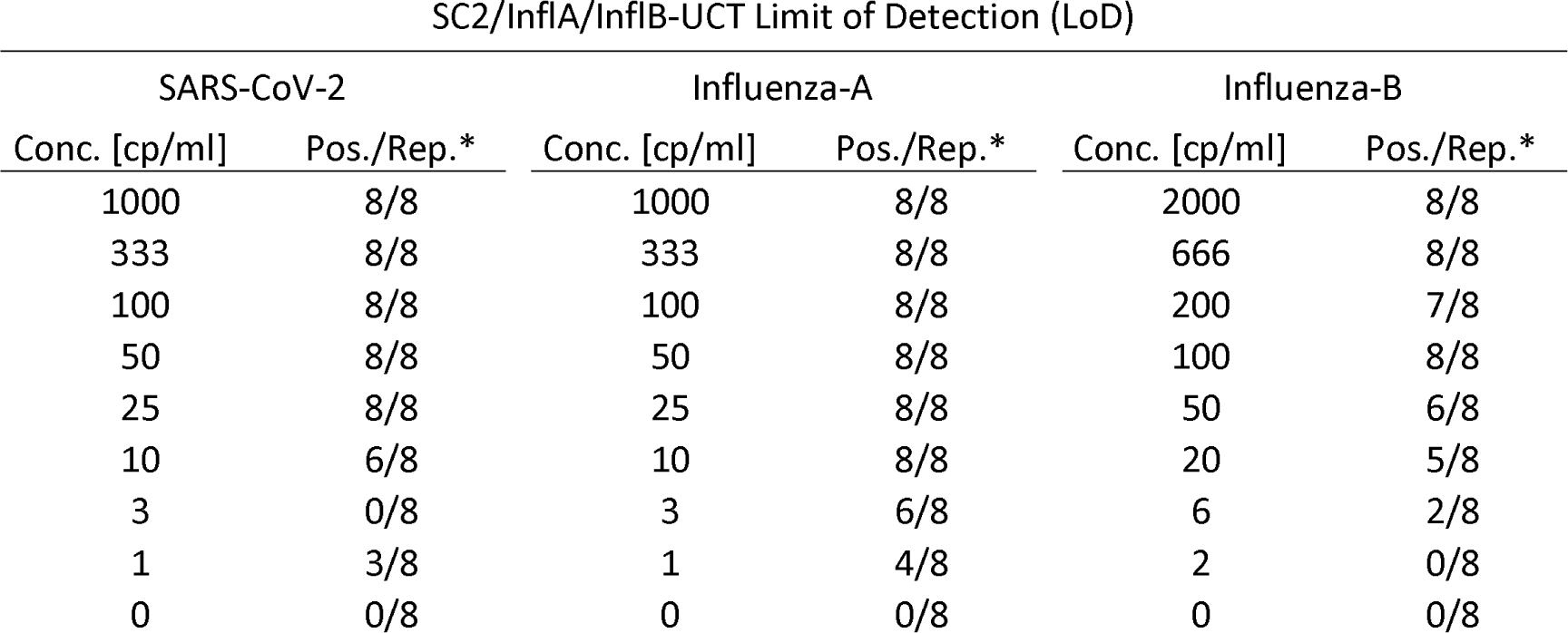
Quantified cell culture stocks and quantified reference material (by Qnostics) was spiked into 1:1 cobas PCR Media in eSwab medium. LOD was determined for all targets simultaneously, meaning that every sample contained the indicated concentrations of each pathogen for each dilution step. * Number of positives / Number of total repeats

**Table 4:** Inclusivity and cross-reactivity. A panel of clinical samples containing respiratory pathogens, as well as relevant external quality control panel samples (INSTAND e.V.) were tested with the SC2/InflA/InflB-UCT. No false positives occurred.

| <u>External Quality Control Panel</u> |  |  |  |  |
| --- | --- | --- | --- | --- |
| Species | Number tested | Target: SC2 | Target: InflA | Target: InflB |
| Influenza A H1N1 pdm09 (A/Michigan/45/2015) | 1 | Negative | **Positive** | Negative |
| Influenza A H7N9 (A/Anhui/1/2013) | 1 | Negative | **Positive** | Negative |
| Influenza A H5N8 (A/DE-SH/Reiherente/AR8444/2016) | 1 | Negative | **Positive** | Negative |
| Influenza B Yamagata (B/Phuket/3073/2013) | 1 | Negative | Negative | **Positive** |
| Influenza B Victoria (B/Colorado/06/2017) | 1 | Negative | Negative | **Positive** |
| Human Coronavirus 229E | 1 | Negative | Negative | Negative |
| Human Coronavirus OC43 | 1 | Negative | Negative | Negative |
| MERS Coronavirus | 1 | Negative | Negative | Negative |
| Parainfluenzavirus 2 | 1 | Negative | Negative | Negative |
| <u>Clinical samples</u> |  |  |  |  |
| Species |  | Target: SC2 | Target: InflA | Target: InflB |
| Human Coronavirus HKU1 | 2 | Negative | Negative | Negative |
| Human Coronavirus NL63 | 1 | Negative | Negative | Negative |
| Human Coronavirus OC43 | 1 | Negative | Negative | Negative |
| Boca-virus | 1 | Negative | Negative | Negative |
| Parainfluenzavirus 3 | 1 | Negative | Negative | Negative |
| Human Metapneumovirus | 2 | Negative | Negative | Negative |
| Rhino-/Enterovirus | 3 | Negative | Negative | Negative |
| Respiratory Syncytial Virus | 2 | Negative | Negative | Negative |
| Mycoplasma pneumoniae | 1 | Negative | Negative | Negative |
| Chlamydia pneumoniae | 1 | Negative | Negative | Negative |
| Pneumocystis jirovecii | 1 | Negative | Negative | Negative |

**Figure 1:**
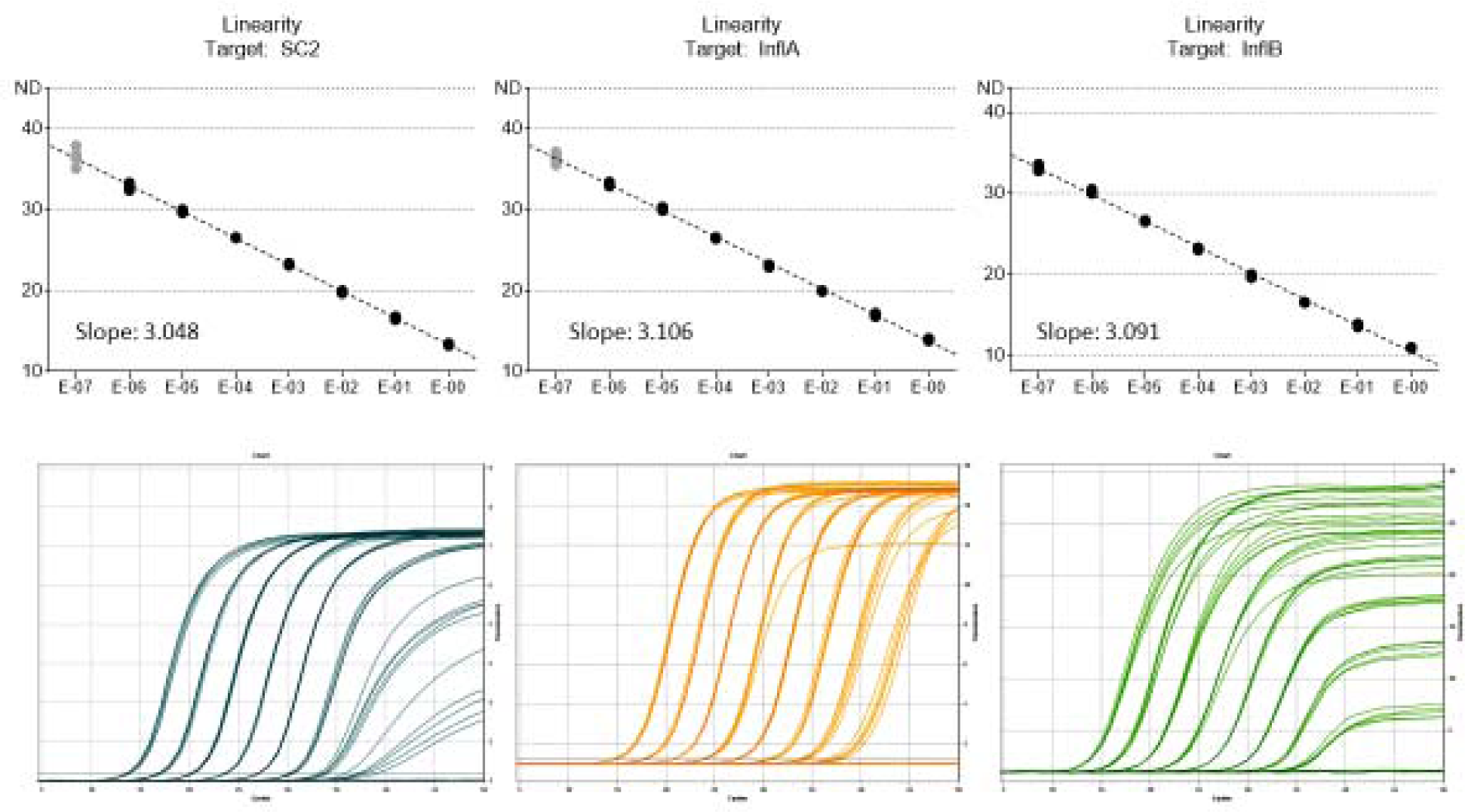
Linearity was determined for all targets simultaneously by serial dilution of SARS-CoV-2 cell culture stocks and Vaxigrip Tetravalent Influenza vaccine in 1:1 cobas PCR Media in eSwab medium. X-axis: dilution factor.

### 3.3 Clinical samples

For clinical evaluation, a total of 164 archived predetermined respiratory swab samples were subjected to the SC2/InflA/InflB-UCT. Clinical samples were oropharyngeal or nasopharyngeal swabs, performed using eSwab sample collection kits (Copan, Italy), containing 1 ml Amies-Medium. 1 – 2 ml of cobas PCR Media (≤⍰40% guanidine hydrochloride in Tris-HCL buffer) were added to samples prior to analysis in routine diagnostics. Samples were stored up to three months at −20°C for SARS-CoV-2 and up to three years for Influenza-A/B.

SARS-CoV-2 samples were included if positive for Target-1 and Target-2 of the SARS-CoV-2 IVD assay. The Cepheid Xpert Xpress Flu/RSV assay was used to resolve discrepant results for Influenza-A/B virus. Samples predetermined positive (by inhouse methods) for Influenza-A or Influenza-B that tested negative in the SC2/InflA/InflB-UCT were included in the study only if tested positive for both Influenza-A targets or positive for Influenza-B in the Xpert Xpress Flu/RSV-assay.

This work was conducted in accordance with §12 of the Hamburg hospital law (§12 HmbKHG). The use of anonymized samples was approved by the ethics committee, Freie und Hansestadt Hamburg, PV5626.

## 4 Results

### 4.1 Analytical performance

Analytical LoD was determined by Probit-Analysis as 94.9 cp/ml (95% CI: 40.5 – 222.0 cp/ml) for SARS-CoV-2, 14.57 cp/ml (95% CI: 6.7 – 31.6 cp/ml) for Influenza-A and 422.3 cp/ml (95% CI: 213.8 – 834.4 cp/ml) for Influenza-B (*table 3*).

The assay showed good linearity for all targets up to a CT value (cycle threshold) of 33 (*Figure 1*).

In cross-reactivity experiments, no false positives occurred. Avian Influenza-A strains H7N9 and H5N8 were correctly detected by the multiplex assay, demonstrating broad coverage (*table 4*). Nonetheless, it is recommended to verify inclusivity of primer/probe sequences as new Influenza-A strains continuously emerge.

### 4.2 Evaluation of clinical performance

Sensitivity in clinical samples containing respective targets was 98.1% for SARS-CoV-2 (52 samples, median CT: 31.99, IQR: 27.21 – 33.96), 97.67% for Influenza-A (43 samples of which 15 were subtyped as H1N1, median CT: 27.80, IQR: 24.85 – 31.1) and 100% for Influenza-B (19 samples, median CT: 29, IQR: 28 - 30), see *table 5*.

**Table 5:** 164 clinical samples were tested in total, predefined positive for SARS-CoV-2 via SARS-CoV-2 IVD test for the cobas6800 system; predetermined positive for Influenza-A or -B by established inhouse methods or Xpert Xpress Influenza/RSV. Invalid rate was 6.4% (invalids not included in the table). Samples were stored at −20°C between 1 and 36 months.

|  |  | Predetermined clinical specimen |  |  |  |
| --- | --- | --- | --- | --- | --- |
|  |  | SARS-CoV2 | Influenza-A | Influenza-B | Negative* |
| SC2/InfIA/InfIB-UCT | SC2 positive | 51/52 | 0/43 | 0/19 | 0/109 |
|  | InfIA positive | 0/52 | 42/43 | 0/19 | 0/118 |
|  | InfIB positive | 0/52 | 0/43 | 19/19 | 0/142 |
|  | Negative | 1/52 | 1/43 | 0/19 | 47/47 |
|  | Total: | 52 | 43 | 19 | 369† |

*\* Total of samples negative for respective target. † Sum of all predetermined negative samples for each individual pathogen. This includes 47 samples negative for all three pathogens.*

CT values showed good correlation with the SARS-CoV-2 IVD, see *figure 2*. For SARS-CoV-2 a single false negative occurred for a sample containing very low amounts of viral RNA, approximately 184 cp/ml. Another false negative occurred for Influenza A in a sample with low viral load (Xpert Flu/RSV, A1 CT: 33, A2 CT: 35)

**Figure 2:**
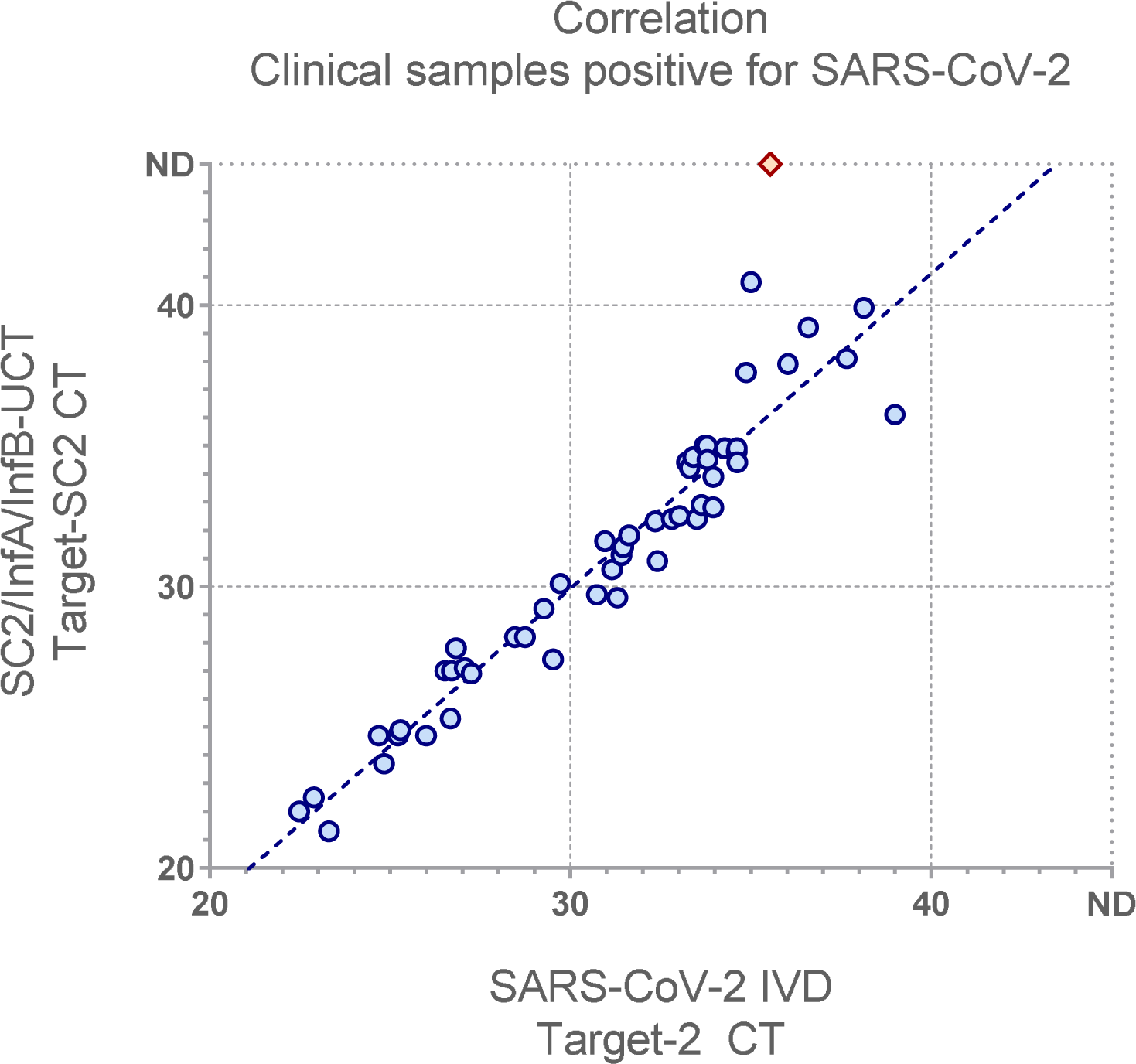
The SC2/InflA/InflB-UCT SC2-CT was compared to the SARS-CoV-2 IVD Target-2 CT (used for quantification and best in regards to linear range) to illustrate correlation with a well-established assay. Red diamond indicates false negative for the multiplex assay. ND = Not detected.

A total of 50 predetermined samples negative for SARS-CoV-2 (by SARS-CoV-2 IVD) and Influenza-A/B (by LDT assay) were subjected to the SC2/InflA/InflB-UCT. No false positives occurred; inhibition rate in eSwab samples was 6.4%.

As a proof of concept, a dilution series of a SARS-CoV-2 positive patient sample was prepared and subjected to testing in the presence and absence of high levels of Influenza-A- and Influenza-B-RNA (*supplement figure 1*). No relevant impact on SARS-CoV-2 detection was observed.

## 5 Discussion

In this study we present a functional SARS-CoV-2/Influenza-A/Influenza-B multiplex assay for the cobas6800 high-throughput platform, featuring analytical and clinical performance comparable to currently used methods for the detection of these pathogens in clinical specimens. The ability to screen for all three viruses in a single reaction allows for streamlined workflows and conservation of resources during the coming Influenza-season. By this time, multiple commercial providers have announced, or are in the process of rolling out multiplex-assays for detection of SARS-CoV-2, Influenza-A/B and RSV. However, these are for the most part designed for a point-of-care rapid testing approach and not high-throughput testing. Moreover, the CDC has itself recently published primer-sets for a SARS-CoV-2/Influenza-A/Influenza-B multiplex assay, which relies on manual PCR workflows (10). However, the ability to employ inhouse assays on fully automated PCR platforms provides vastly improved scalability of testing capacity and less limitations due to availability of trained personnel.

Most currently available commercial SARS-CoV-2 tests with FDA emergency authorization are designed as multi-target assays to account for emerging mutations. While the original Sarbeco-E primerset by Corman et al. (6) resides in a particularly stable region of the SARS-CoV-2 genome, single mutations have been reported within the ever-growing catalogue of available whole genome sequences (13). The RdRp/Hel assay by Chan et al. (7) was modified and adapted as a second target to provide additional security for inclusivity in SARS-CoV-2 detection. Both assays were allocated to the same channel as positivity of any single one would constitute a positive result. There was no indication that presence of Influenza-A and/or Influenza-B RNA within the reaction substantially impairs sensitivity for SARS-CoV-2. If necessary, each SARS-CoV-2 assay can be analysed separately by moving RdRp/Hel detection to channel 1, using the following probe (or comparable): 5’ Atto425-TTAAGATGT(BMN-Q535)GGTGCTTGCATACGTAGAC -BMN-Q535 3’ (*see supplement figure 2*). It has to be acknowledged that the SARS-CoV-2 assays used for this multiplex-setup are technically not specific for SARS-CoV-2 but for the Sarbeco-subgenus of betacoronaviruses, including SARS-CoV (from 2003) and SARS-like bat viruses.

In conclusion, we provide analytical and clinical evaluation of a SARS-CoV-2/Influenza-A/Influenza-B multiplex assay for the cobas6800 high-throughput platform. Performance for each target was comparable to existing solutions currently in use in diagnostic practice. Our novel assay may prove useful for streamlining diagnostics during the upcoming Influenza-season.

## Data Availability

Data available upon request

## 6 Author contribution

Conceptualization: DN, MA, SP, and ML. Methodology and Investigation: DN. Original draft preparation: DN and SP. Review and editing: DN, AH, MA, SP and ML Supervision: SP and ML. All authors agreed to the publication of the final manuscript.

## 8.3 supplement Figure 1

Supplementary Figure 1: A patient sample was used for two 10-fold dilution series in 1:1 cobas PCR Media and eSwab medium, one containing Vaxigrip tetravalent Influenza vaccine in order to rule out relevant interference of high levels of Influenza-RNA for SARS-CoV-2 detection.

## 8.4 supplement Figure 2

Supplement Figure 2: Amplification curves of the SC2/InflA/InflB-UCT using a Atto425 probe for detection of RdRp/Hel on channel one. In this fashion, both SARS-CoV-2 targets can be analyzed separately. This leads to increased cross-talk of the Yakima-Yellow probe into Channel 2 due to lower background fluourescence.

## References

1. Wu Z, McGoogan JM. 2020. Characteristics of and Important Lessons From the Coronavirus Disease 2019 (COVID-19) Outbreak in China: Summary of a Report of 72314 Cases From the Chinese Center for Disease Control and Prevention. JAMA doi:10.1001/jama.2020.2648.

2. Stokes EK, Zambrano LD, Anderson KN, Marder EP, Raz KM, El Burai Felix S, Tie Y, Fullerton KE. 2020. Coronavirus Disease 2019 Case Surveillance - United States, January 22-May 30, 2020. MMWR Morbidity and mortality weekly report 69:759–765.

3. Eigner U, Reucher S, Hefner N, Staffa-Peichl S, Kolb M, Betz U, Holfelder M, Spier G, Pfefferle S, Lutgehetmann M. 2019. Clinical evaluation of multiplex RT-PCR assays for the detection of influenza A/B and respiratory syncytial virus using a high throughput system. J Virol Methods 269:49–54.

4. Pfefferle S, Reucher S, Nörz D, Lütgehetmann M. 2020. Evaluation of a quantitative RT-PCR assay for the detection of the emerging coronavirus SARS-CoV-2 using a high throughput system. Eurosurveillance 25:2000152.

5. Poljak M, Korva M, Knap Gašper N, Fujs Komloš K, Sagadin M, Uršič T, Avšič Županc T, Petrovec M. 2020. Clinical evaluation of the cobas SARS-CoV-2 test and a diagnostic platform switch during 48 hours in the midst of the COVID-19 pandemic. Journal of Clinical Microbiology doi:10.1128/jcm.00599-20:JCM.00599-20.

6. Corman VM, Landt O, Kaiser M, Molenkamp R, Meijer A, Chu DK, Bleicker T, Brunink S, Schneider J, Schmidt ML, Mulders DG, Haagmans BL, van der Veer B, van den Brink S, Wijsman L, Goderski G, Romette JL, Ellis J, Zambon M, Peiris M, Goossens H, Reusken C, Koopmans MP, Drosten C. 2020. Detection of 2019 novel coronavirus (2019-nCoV) by real-time RT-PCR. Euro Surveill 25.

7. Chan JF-W, Yip CC-Y, To KK-W, Tang TH-C, Wong SC-Y, Leung K-H, Fung AY-F, Ng AC-K, Zou Z, Tsoi H-W, Choi GK-Y, Tam AR, Cheng VC-C, Chan K-H, Tsang OT-Y, Yuen K-Y. 2020. Improved Molecular Diagnosis of COVID-19 by the Novel, Highly Sensitive and Specific COVID-19-RdRp/Hel Real-Time Reverse Transcription-PCR Assay Validated <em>In Vitro</em> and with Clinical Specimens. Journal of Clinical Microbiology 58:e00310–20.

8. WHO. 2017. WHO information for the molecular detection of influenza viruses. WHO Website.

9. Terrier O, Josset L, Textoris J, Marcel V, Cartet G, Ferraris O, N’Guyen C, Lina B, Diaz J-J, Bourdon J-C, Rosa-Calatrava M. 2011. Cellular transcriptional profiling in human lung epithelial cells infected by different subtypes of influenza A viruses reveals an overall down-regulation of the host p53 pathway. Virology journal 8:285–285.

10. CDC. 2020. Research Use Only CDC Influenza SARS-CoV-2 (Flu SC2) Multiplex Assay Real-Time RT-PCR Primers and Probes. CDC Website.

11. Pfefferle S, Huang J, Nörz D, Indenbirken D, Lütgehetmann M, Oestereich L, Günther T, Grundhoff A, Aepfelbacher M, Fischer N. 2020. Complete Genome Sequence of a SARS-CoV-2 Strain Isolated in Northern Germany. Microbiology Resource Announcements 9:e00520–20.

12. Nörz D, Frontzek A, Eigner U, Oestereich L, Fischer N, Aepfelbacher M, Pfefferle S, Lütgehetmann M. 2020. Pushing beyond specifications: Evaluation of linearity and clinical performance of a fully automated SARS-CoV-2 RT-PCR assay for reliable quantification in blood and other materials outside recommendations. medRxiv doi:10.1101/2020.05.28.20115469:2020.05.28.20115469.

13. Vogels CBF, Brito AF, Wyllie AL, Fauver JR, Ott IM, Kalinich CC, Petrone ME, Casanovas-Massana A, Catherine Muenker M, Moore AJ, Klein J, Lu P, Lu-Culligan A, Jiang X, Kim DJ, Kudo E, Mao T, Moriyama M, Oh JE, Park A, Silva J, Song E, Takahashi T, Taura M, Tokuyama M, Venkataraman A, Weizman O-E, Wong P, Yang Y, Cheemarla NR, White EB, Lapidus S, Earnest R, Geng B, Vijayakumar P, Odio C, Fournier J, Bermejo S, Farhadian S, Dela Cruz CS, Iwasaki A, Ko AI, Landry ML, Foxman EF, Grubaugh ND. 2020. Analytical sensitivity and efficiency comparisons of SARS-CoV-2 RT–qPCR primer–probe sets. Nature Microbiology doi:10.1038/s41564-020-0761-6.

