## Supplement Figures for "Clinical evaluation of a fully automated, lab developed multiplex RT-PCR assay integrating dual-target SARS-CoV-2 and Influenza–A/B detection on a high-throughput platform"

### Slide 1
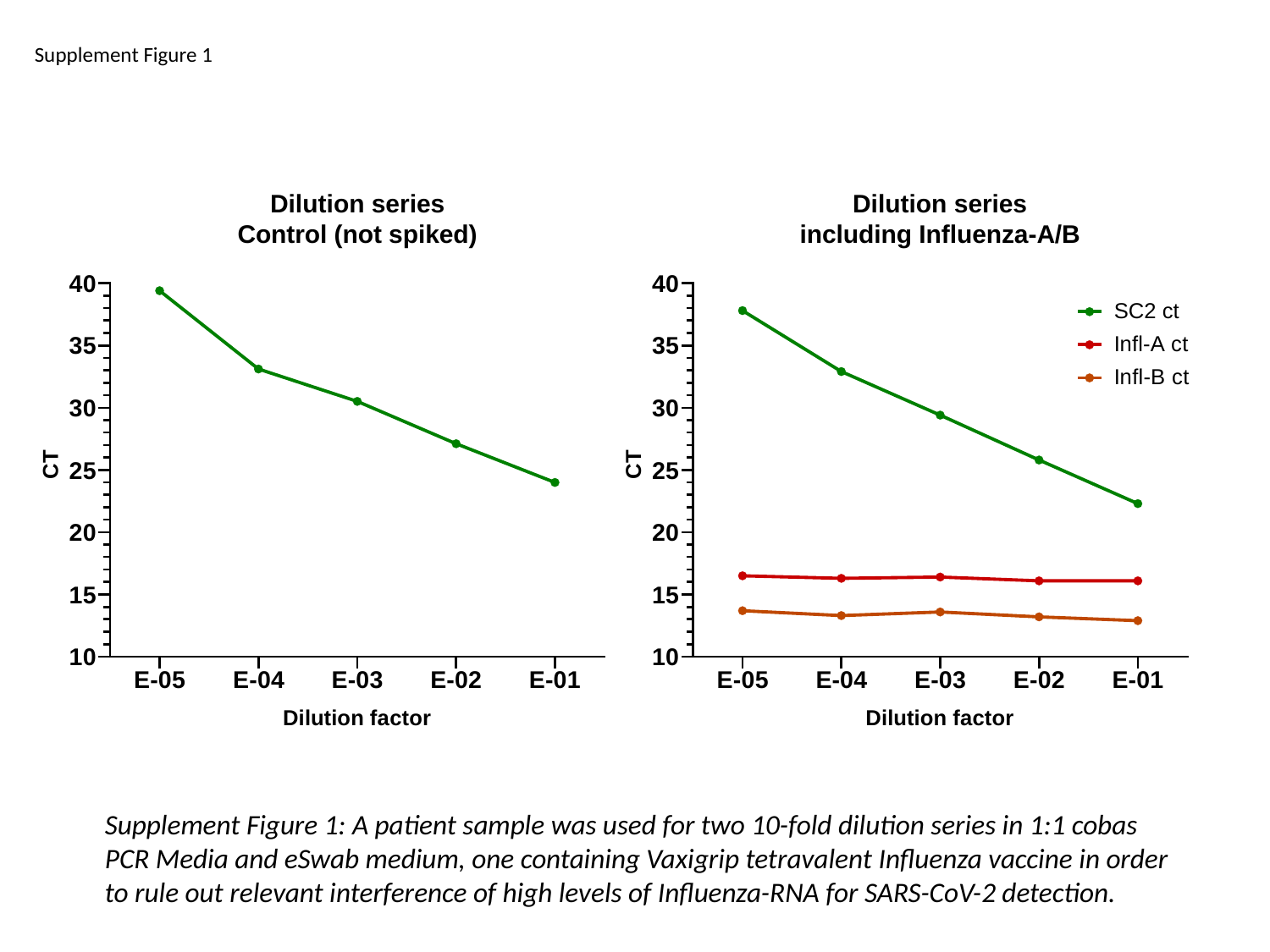

Supplement Figure 1
Supplement Figure 1: A patient sample was used for two 10-fold dilution series in 1:1 cobas PCR Media and eSwab medium, one containing Vaxigrip tetravalent Influenza vaccine in order to rule out relevant interference of high levels of Influenza-RNA for SARS-CoV-2 detection.

### Slide 2
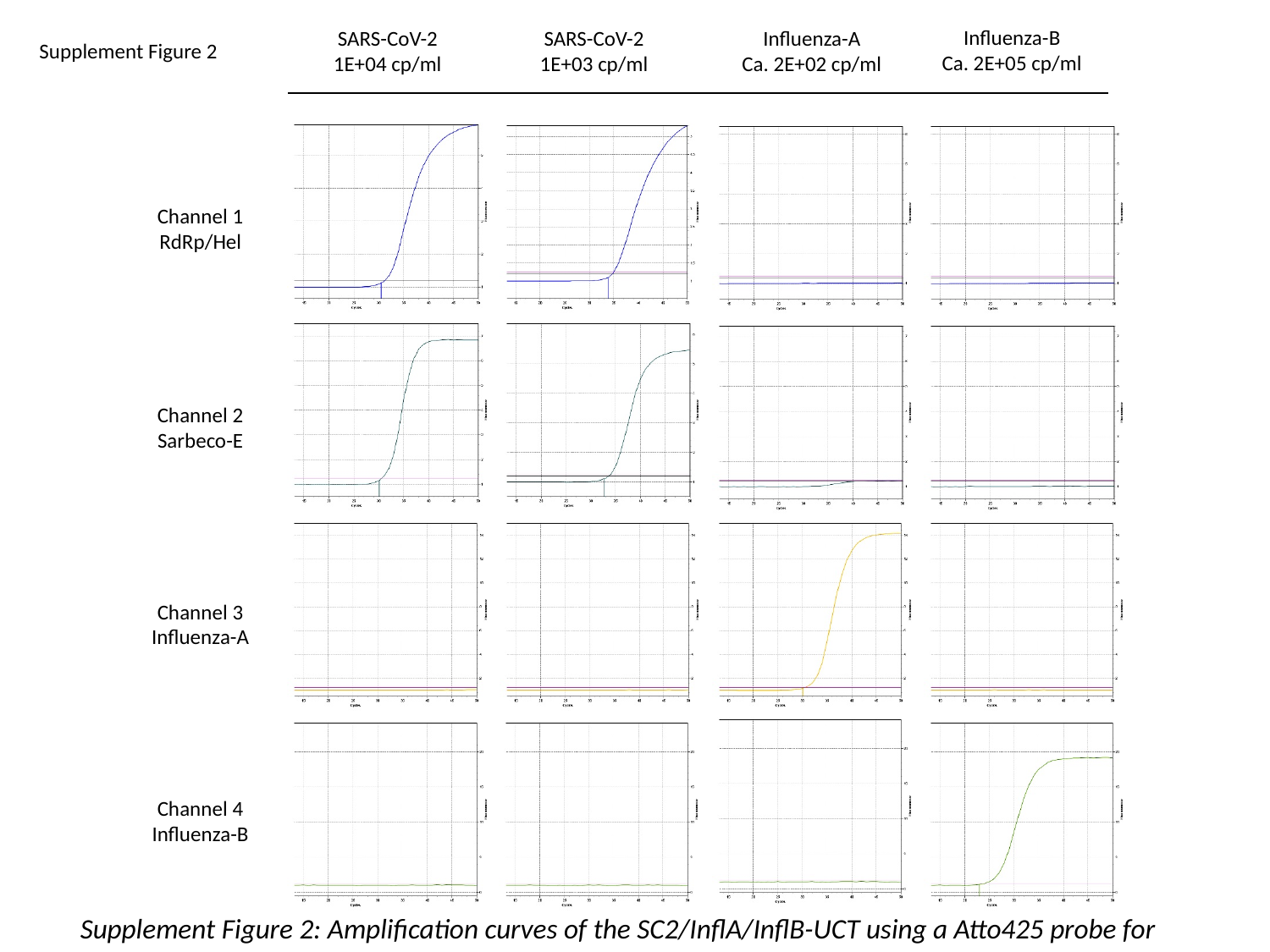

Influenza-B
Ca. 2E+05 cp/ml
Influenza-A
Ca. 2E+02 cp/ml
SARS-CoV-2
1E+04 cp/ml
SARS-CoV-2
1E+03 cp/ml
Supplement Figure 2
Channel 1
RdRp/Hel
Channel 2
Sarbeco-E
Channel 3
Influenza-A
Channel 4
Influenza-B
Supplement Figure 2: Amplification curves of the SC2/InflA/InflB-UCT using a Atto425 probe for detection of RdRp/Hel on channel one. In this fashion, both SARS-CoV-2 targets can be analyzed separately. This leads to increased cross-talk of the Yakima-Yellow probe into Channel 2 due to lower background fluourescence.
